# The prevalence and factors associated with alcohol, cigarette, and marijuana use among adolescents in 25 African countries: evidence from Global School-Based Health Surveys

**DOI:** 10.1101/2024.05.21.24307637

**Authors:** Retselisitsoe Pokothoane, Terefe Gelibo Agerfa, Josiane Djiofack Tsague, Noreen Dadirai Mdege

**Affiliations:** Development Gateway: an IREX Venture, Washington DC, United States; Research Unit on the Economics of Excisable Products (REEP), School of Economics, University of Cape Town, Cape Town, South Africa; Public Health, ICAP at Columbia University Mailman School of Public Health, Addis Ababa, Ethiopia; Department of Health Sciences, University of York, York, United Kingdom; Centre for Research in Health and Development, York, United Kingdom

## Abstract

**Objectives:** To provide first comprehensive estimates of the prevalence of psychoactive substances: alcohol, cigarettes and marijuana and their associated factors among school-going adolescents in 25 African countries, and thereby contribute to the evidence base of substance use in Africa.

**Methods:** We use the publicly available Global School-Based Health Survey (GSHS) data from 25 African countries collected between 2003 and 2017. We used descriptive statistics to estimate the prevalence of alcohol, cigarette, and marijuana use as well as their dual use among adolescents aged 11 – 16 years. Additionally, we used logistic regressions to model factors associated with the use of each substance, with adjusted odds ratios (ORs) and their 95% confidence intervals (CIs) as the measures of association.

**Results:** The prevalence of alcohol use among African adolescents was 10.6% [95% CI = 9.6, 11.8], that for cigarette smoking was 6.9% [95% CI: 6.1, 7.8], and it was 3.8% [95% CI: 3.2, 4.4] for marijuana. The prevalence of dual use of alcohol and cigarettes was 1.5% [95% CI: 1.2, 1.9], that of alcohol and marijuana was 0.9% [95% CI: 0.7, 1.1], and it was 0.8% [95% CI: 0.6, 1.0] for cigarettes and marijuana. The prevalence of cigarette smoking was significantly higher among boys than girls, but that of alcohol and marijuana was insignificant. The use of alcohol, cigarettes, or marijuana was associated with the West African region, higher-income country group, having parents that smoke any tobacco products, being bullied, missing classes without permission, and experiencing sadness and hopelessness in the previous month were positively associated with being a current user of these products.

**Conclusions:** Africa should invest in data collection on substance use among adolescents who are in and out of school. At both primary and secondary school levels, African countries should develop mentorship and other interventions that fuel positivity and discourage bad practices among students to ultimately reduce substance use.

**WHAT IS ALREADY KNOWN ON THIS TOPIC:**

➢ At the country level, geography, predominant religion, and income level are risk factors for substance use.
➢ At the individual level, home environment, being bullied, feeling sad and hopeless, and having suicidal thoughts are positively associated with students’ substance use in African primary and secondary schools.

**WHAT THIS STUDY ADDS:**

➢ In Africa, cigarette smoking among adolescents differs significantly by gender across different age groups. Nonetheless, for alcohol and marijuana use, there is no statistically significant difference by gender across age groups.
➢ The common dual use of unhealthy products among adolescents is in the form of alcohol and cigarettes.
➢ Staying in the West African region and missing primary or secondary school classes increases the chances of consuming alcohol, cigarettes, and marijuana in both single and dual use.

**HOW THIS STUDY MIGHT AFFECT RESEARCH, PRACTICE OR POLICY:**

➢ School-based interventions should be developed or further improved to fuel positivity among students and, finally, minimize negative emotions and activities that lead to substance use.

## INTRODUCTION

Psychoactive substance use (hereafter referred to as substance use) is one of the primary risky behaviours among adolescents, and it negatively affects their health, both in the current and long-term periods.^1^ Globally, the common substances consumed during adolescence are alcohol, tobacco, and marijuana (cannabis).^2^ The harmful use of alcohol consumption is one of the leading risk factors for public health and is responsible for around 3 million deaths annually.^3^ Similarly, tobacco smoking is addictive and kills over 8 million smokers globally.^4^ Marijuana is known to be associated with mental health problems among adolescents, which could persist in their adulthood^5^ and ultimately become a threat to their human capital development.

The United Nations (UN) Sustainable Development Goals (SDG) 3.5 requires UN member states to strengthen the control and prevention of substance use to promote good health and well-being of everyone at all ages.^6^ Given that substance use typically begins during adolescence,^1^ to control the use of these substances, it is essential to understand their prevalence of use in this age group. In Africa, even though substance use continues to be a major public health concern, the prevalence of the use of different substances is poorly documented.^7^ Among the World Health Organization (WHO) member states, the standard data source for monitoring substance use beyond tobacco among adolescents is the Global School-Based Health Based Survey (GSHS). However, so far, cross-country evidence that has analyzed and synthesized these data sets has been limited to a few African countries^2 5 8 9^ and restricted specific age groups that often exclude early adolescents (e.g., 11 -12-year-olds) or specific products.

For the first time, we provide more comprehensive estimates of the prevalence of cigarettes, alcohol, and marijuana and their dual use with each other, covering 25 African countries that have conducted at least one wave of GSHS. The age range available in the datasets varied across these 25 African countries. Thus, for this study, we focused on the 11 -16 years age group as this was the age range that was common across the datasets, allowing us to include early adolescents. We also assessed the country-level and individual-level factors associated with the prevalence of the single and dual use of these substances.

## METHODS

### Data sources

This study used the latest waves of the GSHS data from the 25 African countries that have conducted at least one round of the survey. The datasets cover the period 2003 to 2017. GSHS is a school-based survey conducted among adolescents in grades that correspond with the age group 13 – 17 years in WHO member states. Nonetheless, the GSHS surveys in many countries tend to include adolescents aged 11 - 18 years. The overall objective of the survey is to collect information on the leading causes of morbidity and mortality among adolescents, including substance use. The survey was developed by WHO in collaboration with Centres for Diseases Control and Prevention (CDC). GSHS is a two-stage national cluster sample survey in which a sample of schools was selected in the first stage. In the second stage, classes were selected randomly within the selected schools, and all students within the selected classrooms participated in the self-administered questionnaire.^10^

Most of the 25 African GSHS surveys included participants aged 11 – 16 years, except in 8 countries: Benin, Eswatini, Ghana, Liberia, Morocco, Mozambique, Mauritius, and Namibia, which included participants outside this age group (i.e., 17 – 18 years).To ensure uniformity across all surveys, we excluded any participants outside the 11 – 16 years age group in the analysis.

### Substance use variables

Outcome variables based on questions asked in GSHS and were defined as follows:

#### Current alcohol use

consuming at least one drink containing alcohol for at least a day in the past 30 days before the survey. This was obtained from the question, “During the past 30 days, on how many days did you have at least a standard drink containing alcohol?” Those who reported at least one day were considered as current alcohol drinkers, and those who did not report any smoking day were considered as current non-drinkers.

#### Current cigarette smoking

smoking cigarettes for at least one day in the past 30 days preceding the survey. The participants were asked, “During the past 30 days, how many days did you use cigarettes?” Those who reported at least one day were classified as current cigarette smokers. Otherwise, they were classified as non-cigarette smokers.

#### Current marijuana use

using marijuana at least once a month. This was retrieved from the following survey question, “During the past 30 days, how many times did you use marijuana?” Those who reported using marijuana at least once in 30 days were categorized as current marijuana smokers, and those without any time were considered non-current users.

#### Current dual alcohol and cigarette use

Participants who reported using alcohol and cigarettes in the past 30 days were categorized as dual users. Those who used none or only one of the products were considered non-dual users.

#### Current dual use of alcohol and marijuana

Participants who reported using alcohol and marijuana in the past 30 days were classified as current dual users of alcohol and marijuana. Those who reported using none of these products or just one of them but not both were classified as non-dual users.

#### Current dual use of cigarettes and marijuana

Participants who reported using both cigarettes and marijuana in the past 30 days were classified as dual cigarette and marijuana users. Those who reported using none of these substances or using only one of them were considered non-dual users.

### Other variables of interest

Using information from the literature^2 5 7 8^ and the GSHS questionnaires, our analysis also included individual-level (demographic and interpersonal) and country-level (i.e., geographic), as indicated below.

#### Demographic variables

The demographic variables include gender, age, number of close friends, parental/guardian smoking, and current school-level (primary/secondary).^2 5 7 8^ The current school-level variable was constructed using the information on students’ current class grades. We could not use the ‘grade’ variable directly as it is because African countries have different education systems and, hence, different grade names. The choice of the mentioned variables was due to their association with the use of different substances in the literature. Because of data constraints, we could not capture other important factors, such as individual income (pocket money) and product prices.

#### Interpersonal factors

These factors were chosen on the basis of their association with the products in question from different studies.^2 8^ We use the following personal/interpersonal variables:

##### Being sad and hopeless

This variable assessed students’ stress-oriented feelings. It is a dummy variable generated from the survey question, “During the past 12 months, did you ever feel so sad or hopeless almost every day for two weeks or more in a row that you stopped doing your usual activities?” The variable assumes 1 if a student responded “yes” and 0 if they responded “no”.

##### Bullying

This variable represented violence and unintentional injury behaviour among students. It is a dummy variable generated from the survey question, “During the past 30 days, on how many days were you bullied?” This variable equals to 1 if a respondent reported at least one day of experiencing bullying and 0 otherwise.

##### Missing school

This represents students’ experiences at school. It is a dummy variable generated from the survey question, “During the past 30 days, on how many days did you miss classes or school without permission?” This variable equals to 1 if a respondent reported at least one day of missing school without permission, and 0 otherwise.

#### Country-level variables

Geographic location, country wealth, level of development, and religion have been found to be associated with substance use.^2 8^ Thus, we use 4 types of country-level (geographic and wealth-related) variables.

##### Africa region

This categorical variable classifies African countries by region: East Africa, North Africa, Southern Africa, and West Africa.^11^ Central Africa region is not represented in this study since none of the countries in that region have GSHS.

##### World Bank income group

This categorical variable classifies countries into low-income, lower-middle-income, and upper-middle-income groups. It is obtained from the 2023 World Bank development indicators.^12^

##### Religion

This categorical variable assumes the following outcomes: Christian and Muslim (inclusive of Hindu). It represents the dominant religion for each country. Following de la Torre-Luque et al.,^2^ we obtained it from the report entitled “Future of World Religions: Population Growth Projections: 2010 – 2050”.^13^

##### Urbanization

This continuous variable represents the percentage of people living in the urban areas for each country and is obtained from the United Nations World Urbanization prospects.^14^

## Data Analysis

We used Stata version 17^15^ and appended all 25 surveys to obtain the consolidated sample of 66,145 adolescents aged 11 -16 years. We used the dataset sample weights to adjust the surveys for non-responses and sample representation in each country. We obtained pooled prevalence estimates of the three substances for all 25 countries. Prevalence estimates were computed for each country and by African region, World Bank income group, and demographic groups (i.e., age and gender). All prevalence estimates were reported with the 95% Confidence Intervals (CI’s).

Multivariate logistic regression analysis was used to identify the factors associated with the use of alcohol, cigarettes, marijuana, dual alcohol and cigarettes, dual alcohol and marijuana, and dual cigarettes and marijuana. We estimated six independent logistic regression models differing in terms of the dependent variables, but similar in terms of independent variables. We presented the adjusted Odds Ratios (OR), their 95% CI’s, and the associated p-values.

## RESULTS

### Sample characteristics

Table 1 presents the descriptive statistics. The mean age in the sample was 14.3 years (standard deviation (sd) = 2.6 years). The sample comprised more girls (53.3%) than boys (47.7%). Most of the students were in secondary schools and resided in African countries with Christianity as the dominant religion, which were also in the lower-middle-income group. Close to 30% of the sample came from East, North, and Southern Africa. We did not identify any GSHS dataset from countries in Central Africa (Table 1). The mean percentage of people living in the urban areas (urbanization) was 48.8% (sd =7.8%). The respondents in the GSHS predominantly (30%) came from the survey period 2003 – 2005. Close to half (46.3%) reported experiencing bullying at least one day during the past 30 days, and this was 45.1% for missing class without permission and 28.1% for ever feeling sad and hopeless.

**Table 1.** Characteristics of the participants from the GSHS in 25 African countries.

| Variable | Number of observations (n) | Percent (%) |
| --- | --- | --- |
| **All** | 66,145 | 100% |
| **Gender** |  |  |
| Boys | 31, 055 | 47.7% |
| Girls | 34, 006 | 53.3% |
| **Age** |  |  |
| 11 years | 1,374 | 2.1% |
| 12 years | 4,759 | 7.3% |
| 13 years | 11,786 | 18.1% |
| 14 years | 15,250 | 23.4% |
| 15 years | 16,807 | 25.8% |
| 16 years | 15,081 | 23.2% |
| **School Level** |  |  |
| Primary | 12,055 | 18.6% |
| Secondary | 52, 933 | 81.4% |
| **Region** |  |  |
| East Africa | 19,273 | 29.1% |
| North Africa | 19,614 | 29.7% |
| Southern Africa | 18,164 | 27.5% |
| West Africa | 9,094 | 13.8% |
| **World Bank Income Group** |  |  |
| Upper middle income | 12,223 | 18.5% |
| Lower middle income | 42,107 | 63.7% |
| Low income | 11,815 | 17.8% |
| **Predominant Country Religion** |  |  |
| Christian | 34, 921 | 52.8% |
| Hindu | 2,538 | 3.8% |
| Muslim | 28, 686 | 43.4% |
| **Survey year** |  |  |
| 2003 – 2005 | 20,046 | 30.3% |
| 2006 – 2008 | 6,889 | 10.4% |
| 2009 – 2011 | 11,522 | 17.4% |
| 2012 – 2014 | 12,084 | 18.3% |
| 2015 – 2017 | 15,604 | 23.6% |
| **Bullying** |  |  |
| Yes | 24,663 | 46.3% |
| No | 28,618 | 53.7% |
| **Missed classes without permission** |  |  |
| Yes | 26,997 | 45.1% |
| No | 32,837 | 54.9% |
| **Feeling sad and hopeless** |  |  |
| Yes | 13,609 | 28.1% |
| No | 34,852 | 71.9% |

### Prevalence of current use of alcohol, cigarettes, and marijuana

Among the three substances, being a current alcohol drinker was more common (10.6%, [95% CI = 9.6, 11.8]), followed by being a current cigarette smoker (6.9%, [95% CI = 6.1, 7.8]), and current marijuana user (3.8%, [95% CI = 3.2, 4.4]). In terms of dual use, the prevalence of current dual use of alcohol and cigarettes was 1.5% [95% CI = 1.2, 1.9], and those for dual use of alcohol and marijuana, and cigarettes and marijuana were similar, 0.9% [95% CI = 0.7, 1.1] and 0.8% [95% CI = 0.6, 1.0], respectively (Table 2). Figure 1 shows the prevalence of cigarettes, alcohol, and marijuana use by gender and age. For those aged 12 – 16 years, there was a statistically significant difference in the prevalence of cigarette smoking by gender, but not among those aged 11 years. For the prevalence of alcohol and marijuana use, there is no statistically significant difference by gender for all age groups.

**Table 2.** Prevalence of alcohol, cigarettes, and marijuana use by country, region, and income group.

|  | Alcohol (95% CI) | Cigarettes (95% CI) | Marijuana (95% CI) | Dual alcohol & cigarette use (95% CI) | Dual alcohol & marijuana use (95% CI) | Dual cigarette & marijuana use (95% CI) |
| --- | --- | --- | --- | --- | --- | --- |
| **East Africa** |  |  |  |  |  |  |
| Djibouti (2007) | X | 6.1% (4.8, 7.8) | X | X | X | X |
| Kenya (2003) | 12.9% (9.1, 18.1) | 16.1% (12.2, 21.1) | X | 6.9% (4.0, 11.7) | X | X |
| Mauritius (2017) | 23.4% (21.0, 26.1) | 16.5% (13.7, 19.8) | 5.2% (3.8, 7.2) | 9.6% (7.9, 11.6) | 3.6% (2.5, 5.0) | 3.8% (2.7, 5.4) |
| Seychelles (2015) | 47.6% (44.6, 50.7) | 19.7% (17.4, 22.2) | 9.3% (7.5, 11.4) | 15.4% (13.3, 17.7) | 6.6% (5.3, 8.3) | 6.2% (5.0, 7.8) |
| Sudan (2012) | X | 6.8% (5.1, 8.9) | X | X | X | X |
| Tanzania (2014) | 4.6% (3.5, 6.0) | 4.7% (3.6, 6.1) | 2.6% (1.9, 3.6) | 1.4% (0.9, 2.2) | 1.3% (0.8, 1.9) | 0.9% (0.6, 1.3) |
| Uganda (2003) | 10.7% (9.1, 12.4) | 4.9% (3.8, 6.4) | X | 2.2% (1.6, 2.9) | X | X |
| **Regional prevalence** | **7.6% (6.2, 9.3)** | **7.9% (6.7, 9.4)** | **2.7% (2.0, 3.7)** | **2.7% (1.9, 3.9)** | **0.9% (0.7, 1.3)** | **0.6% (0.4, 0.8)** |
| **North Africa** |  |  |  |  |  |  |
| Algeria (2011) | X | 9.5% (7.8, 11.6) | 6.9% (3.9, 12.1) | X | X | 1.2% (0.7, 2.1) |
| Egypt (2011) | X | 5.2% (3.1, 8.6) | X | X | X | X |
| Libya (2007) | X | 4.0% (3.3, 4.9) | X | X | X | X |
| Mauritania (2010) | X | 17.5% (13.8, 22.1) | 6.9% (3.9, 12.1) | X | X | 4.5% (2.5, 8.0) |
| Morocco (2016) | X | 5.9% (5.0, 7.0) | 5.4% (4.1, 7.1) | X | X | 2.0% (1.5, 2.7) |
| Tunisia (2008) | X | 8.3% (7.0, 9.8) | X | X | X | X |
| **Regional prevalence** | **X** | **6.4% (5.1, 8.1)** | **3.9% (3.1, 4.8)** | **X** | **X** | **0.7% (0.6, 1.0)** |
| **Southern Africa** |  |  |  |  |  |  |
| Botswana (2005) | 22.5% (20.1, 25.0) | 7.8% (6.7, 9.0) | X | 4.8% (4.1, 5.7) | X | X |
| Eswatini (2013) | X | X | 3.1% (2.1, 4.3) | X | X | X |
| Malawi (2009) | 4.2% (2.0, 8.6) | 4.7% (3.1, 7.1) | X | 1.5% (0.8, 3.0) | X | X |
| Mozambique (2015) | 10.2% (7.0, 14.7) | 2.2% (0.9, 5.0) | 1.4% (0.7, 2.6) | 0.8% (0.4, 1.7) | 0.9% (0.3, 2.4) | 0.3% (0, 1.1) |
| Namibia (2013) | 26.9% (24.7, 29.2) | 8.7% (7.1, 10.7) | 5.1% (3.8, 6.7) | 5.8% (4.8, 7.1) | 3.0% (2.3, 4.1) | 2.6% (1.8, 3.7) |
| Zambia (2004) | 27.1% (24.3, 30.0) | X | X | X | X | X |
| Zimbabwe (2003) | 14.4% (11.8, 17.5) | 9.9% (8.7, 11.3) | X | 4.5% (3.6, 5.7) | X | X |
| **Regional Prevalence** | **12.6% (10.8, 14.6)** | **4.9% (3.9, 6.2)** | **2.2% (1.5, 3.1)** | **2.0% (1.5, 2.5)** | **0.5% (0.3, 0.9)** | **0.3% (0.2, 0.5)** |
| **West Africa** |  |  |  |  |  |  |
| Benin (2016) | 40.0% (34.7, 45.4) | 4.2% (2.7, 6.4) | 1.0% (0.4, 2.4) | 3.8% (2.5, 5.6) | 0.8% (0.3, 2.4) | 0.5% (0.2, 1.4) |
| Ghana (2017) | 14.5% (12.4, 17.0) | 8.5% (5.7, 12.4) | 7.4% (5.1, 10.5) | 4.7% (3.2, 6.9) | 4.1% (3.0, 5.7) | 3.1% (1.9, 5.3) |
| Liberia (2008) | 18.4% (15.5, 21.7) | 7.4% (5.5, 9.8) | 6.3% (4.0, 9.7) | 2.7% (1.7, 4.4) | 2.6% (1.6, 4.2) | 2.7% (1.7, 4.2) |
| Senegal (2020) | 5.7% (3.3, 9.9) | 8.5% (4.7, 15.0) | X | 2.6% (1.3, 5.4) | X | X |
| Sierra Leone (2017) | 11.9% (8.8, 15.8) | X | 4.2% (2.7, 6.6) | X | 2.5% (1.4, 4.3) | X |
| **Regional Prevalence** | **15.3% (13.7, 17.2)** | **7.9% (5.9, 10.5)** | **6.0% (4.4, 8.1)** | **3.7% (2.8, 4.9)** | **2.7% (2.0, 3.6)** | **1.9% (1.2, 3.0)** |
| **WB income group** |  |  |  |  |  |  |
| Upper middle-income | 25.1% (23.7, 26.6) | 6.0% (5.3, 6.8) | 5.2% (1.8, 3.6) | 2.1% (1.8, 2.5) | 2.4% (1.9, 3.0) | 0.7% (0.5, 0.9) |
| Lower middle income | 10.5% (9.2, 11.9) | 7.3% (6.3, 8.4) | 3.8% (3.2, 4.5) | 1.5% (1.2, 2.0) | 0.9% (0.7, 1.2) | 0.9% (0.7, 1.1) |
| Low income | 8.8% (7.3, 10.6) | 4.9% (4.1, 6.0) | 2.6% (1.8, 3.6) | 1.0% (0.7, 1.3) | 0.7% (0.4, 1.1) | 0.2% (0, 0.3) |
| **Overall Prevalence** | **10.6% (9.6, 11.8)** | **6.9% (6.1, 7.8)** | **3.8% (3.2, 4.4)** | **1.5% (1.2, 1.9)** | **0.9% (0.7, 1.1)** | **0.8% (0.6, 1.0)** |

*Notes:* X indicates that the data were not available.

**Figure 1.**
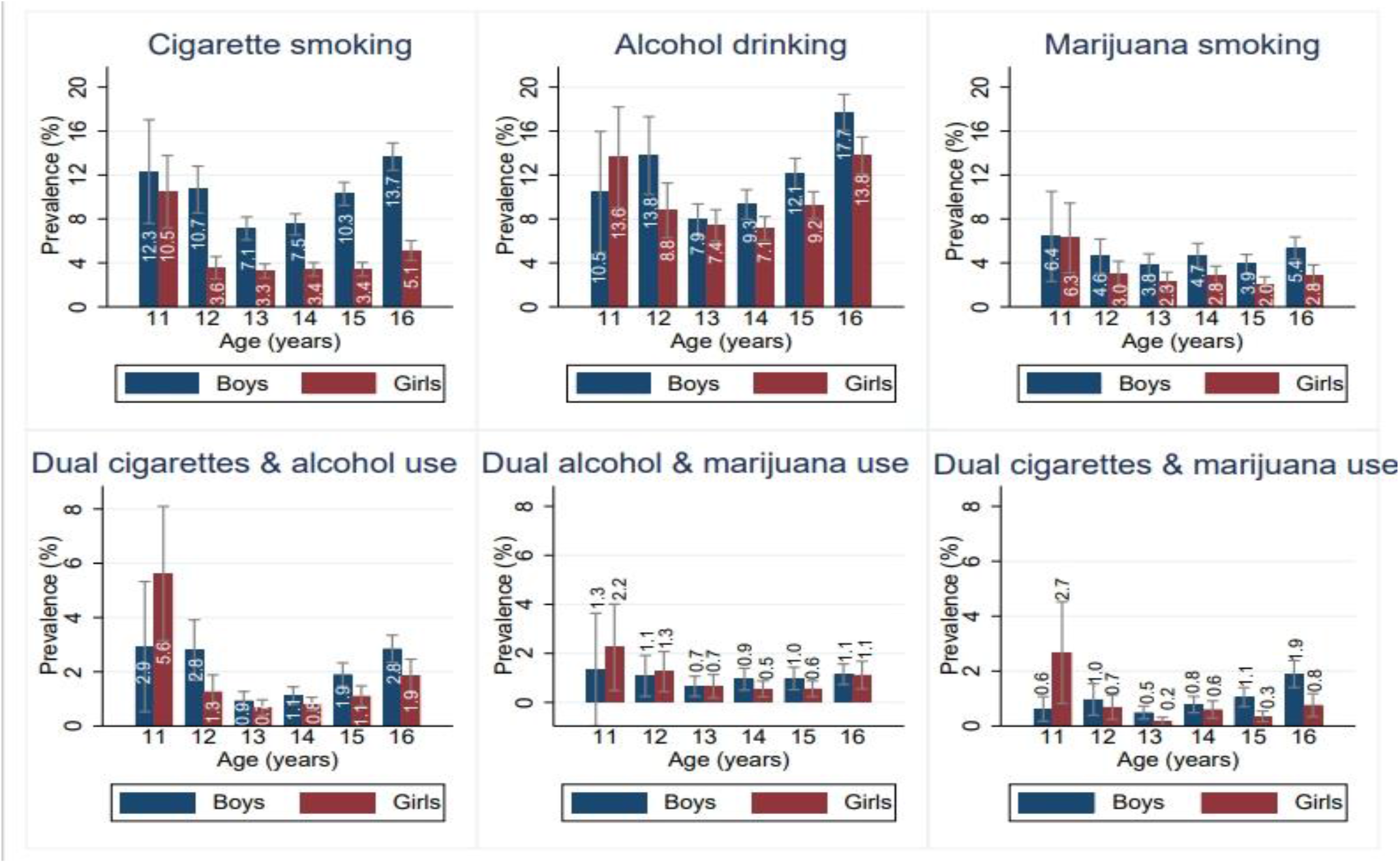
Prevalence of alcohol, cigarettes, and marijuana use: by age and gender

Regionally, the prevalence of alcohol drinking was 15.3% [95% CI: 10.8, 14.6] for West Africa, 12.6 [95% CI: 10.8, 14.6] for Southern Africa, and 7.6% [95% CI: 6.2, 9.3] for East Africa (Table 2). At the country level, the prevalence of alcohol drinking ranged from 4.2% [95% CI = ] (Malawi) to 47.6% (Seychelles). The prevalence of alcohol drinking among adolescents increased with the country’s income group. Gender-wise, this was higher for boys (11.9%, [95% CI = 10.7, 13.2]) than girls (9.2%, [95% CI = 8.1, 10.5]) (Supplementary Table 2).

Cigarette smoking was equally prevalent in East Africa (7.9%, [95% CI: 6.7, 9.4]) and West Africa (7.9, [95% CI: 5.9, 10.5]), and lowest in Southern Africa (4.9%, [95% CI: 3.9, 6.2]) (Table 2). Country-wise, this was highest in Seychelles (24.5%) [95% CI: 21.3, 27.9] and lowest in Mozambique (1.2%) [95% CI: 0.5, 3.0] (Supplementary Table 2). The prevalence of cigarette smoking increased with the country’s World Bank income group. Cigarette smoking was more prevalent among boys (9.8%, [95% CI = 8.4, 11.2]) than girls (3.9%, [95% CI = 3.3, 4.6]). The overall prevalence of marijuana was highest in West Africa, 6.0% [95% CI: 4.4, 8.1], and lowest in Southern Africa, 2.0% [95% CI: 1.5, 3.1]. Gender-wise, this prevalence was higher among boys, 4.6% [95% CI = 3.9, 5.3] compared to girls, 2.7% [95% CI: 2.2, 3.4] (Supplementary Table 2).

The dual use of alcohol and cigarettes was more common than that of alcohol and marijuana or cigarette and marijuana. West Africa had the highest prevalence of the dual use of all the assessed products, ranging from 3.7% [95% CI: 2.8, 4.9] of the concurrent use of alcohol and cigarettes to 1.9% [95% CI: 1.2, 3.0] of the use of both cigarettes and marijuana. The prevalence of dual alcohol and cigarettes ranges from 0.8% [95% CI: 0.4, 1.7) in Mozambique to 15.4% [95% CI: 13.3, 17.7] in Seychelles; that of alcohol and marijuana ranges from 0.9% [95% CI: 0.3, 2.4] in Mozambique to 6.6% [95% CI: 5.3, 8.3] in Seychelles. The least common dual use, that of cigarettes and marijuana, ranges from 0.3% [95% CI = 0, 1.1] to 6.2% [95% CI = 5.0, 7.8] in Seychelles (Table 2). The prevalence estimates of the gendered dual use are presented in Supplementary Table 3. The prevalence rates for the dual use of all the substances are still higher among boys than girls.

Our analysis (i.e. using surveys from 2003 – 2017) includes data that is old, potentially suggesting that prevalence rates may have changed. We performed sensitivity analysis of the prevalence rates by excluding all surveys conducted before 2010. We find no significant differences in the prevalence rates by substance, gender, Africa region, and World Bank income group (See Supplementary Table 4). Thus, our prevalence rates are robust to the exclusion of the datasets that may be considered outdated.

### Factors associated with the current use of alcohol, cigarettes, and marijuana

Students residing in West Africa were more likely to use alcohol (OR 2.7 [95% CI = 1.8, 4.1]) than those from Central Africa. Similarly, those from countries in the upper middle World Bank income group had higher odds of alcohol use (OR 3.8 [95% CI = 2.7, 5.3]) than those from low-income countries. Students with two or more friends were also more likely to be current alcohol users (OR 1.4, [95% CI = 1.0, 1.9]) than those with no friends. Those who had parents/guardians who used any tobacco products were more likely to use alcohol [OR 2.7, 95% CI (2.1, 3.6)] than those without such parents. Those who had experienced bullying (OR 2.2, [95% CI = 2.1, 3.6]) were more likely to use alcohol compared to those who had not been bullied. Those who reported being sad and hopeless were more likely to drink (OR 1.7, [95% CI = 1.5, 2.0]) compared to those who had not experienced sadness and hopelessness, respectively. Finally, students who missed school without permission were more likely to use alcohol (OR 1.6, [95% CI = 1.3, 1.9]) than those who did not (Table 3).

**Table 3.** Logistic regression results of the factors associated with the current use of different substances.

| Variables | Alcohol<br>Odds Ratio (95% CI) | Cigarettes<br>Odds Ratio (95% CI) | Marijuana<br>Odds Ratio (95% CI) |
| --- | --- | --- | --- |
| Age | 1.0 (0.9, 1.2)<br>[0.569] | 1.0 (0.9, 1.1)<br>[0.478] | 0.9 (0.8, 1.1)<br>[0.405] |
| Female (Ref: Male) | 0.9 (0.7, 1.0)<br>[0.069] | 0.5 (0.4, 0.6)<br>[0.000] | 0.6 (0.5, 0.9)<br>[0.010] |
| Education (Ref: Primary) |  |  |  |
| Secondary | 1.1 (0.8, 1.5)<br>[0.502] | 0.7 (0.5, 1.0)<br>[0.031] | 0.6 (0.3, 1.1)<br>[0.113] |
| Africa Region (Ref: East Africa) |  |  |  |
| North Africa | X<br>X | 0.3 (0.2, 0.4)<br>[0.000] | 1.5 (0.5, 4.5)<br>[0.504] |
| Southern Africa | 1.2 (0.9, 1.7)<br>[0.214] | 0.8 (0.6, 1.1)<br>[0.547] | 0.8 (0.3, 2.5)<br>[0.708] |
| West Africa | 2.7 (1.8, 4.1)<br>[0.000] | 1.7 (1.0, 2.7)<br>[0.038] | 3.8 (1.7, 8.4)<br>[0.001] |
| World Bank income group (Ref: Low income) |  |  |  |
| Lower middle income | 0.9 (0.7, 1.1)<br>[0.239] | 1.9 (1.2, 2.8)<br>[0.002] | 2.1 (0.4, 11.4)<br>[0.387] |
| Upper middle income | 3.8 (2.7, 5.3)<br>[0.000] | 3.1 (1.9, 5.1)<br>[0.000] | 5.7 (2.3, 13.9)<br>[0.000] |
| Religion (Ref: Christian) |  |  |  |
| Muslim & Hindu | 0.9 (0.5, 1.5)<br>[0.729] | 3.2 (1.6, 6.7)<br>[0.002] | 0.7 (0.4, 1.0)<br>[0.051] |
| Having close friends (Ref: None) |  |  |  |
| One | 1.1 (0.8, 1.5)<br>[0.661] | 1.1 (0.9, 1.3)<br>[0.520] | 0.6 (0.4, 0.9)<br>[0.015] |
| At least two | 1.4 (1.0, 1.9)<br>[0.037] | 1.0 (0.8, 1.3)<br>[0.850] | 0.6 (0.4, 1.0)<br>[0.048] |
| Parental smoking (Ref: No) | 2.7 (2.1, 3.6)<br>[0.000] | 2.8 (2.3, 3.4)<br>[0.000] | 3.9 (2.8, 5.4)<br>[0.000] |
| Bullying (Ref: No) | 2.2 (2.1, 3.6)<br>[0.000] | 3.0 (2.5, 3.6)<br>[0.000] | 2.6 (2.0, 3.4)<br>[0.000] |
| Sad & hopeless (Ref: No) | 1.7 (1.5, 2.0)<br>[0.000] | 2.4 (2.0, 2.9)<br>[0.000] | 3.1 (2.3, 4.2)<br>[0.000] |
| Missing school (Ref: No) | 1.6 (1.3, 1.9)<br>[0.000] | 1.7 (1.5, 2.0)<br>[0.000] | 2.4 (1.8, 3.3)<br>[0.000] |
| Observations | 21,545 | 30,754 | 17,919 |

*Notes:* The p-values are reported in square brackets. The regression results were weighted, and the standard errors have been adjusted for clustering. The survey year fixed effects have been included in all three regressions. X indicates that the data was unavailable. Urbanization has been dropped due to multicollinearity.

For cigarette smoking, being a female was associated with lower odds of smoking (OR 0.5, [95% CI = 0.4, 0.6]) than being a male. Similarly, secondary school students were less likely to smoke cigarettes (OR 0.7, [95% CI = 0.5, 1.0]) than primary school students. Those who reside in North Africa were less likely to smoke (OR 0.3, [95% CI = 0.2, 0.4]), while those who reside in West Africa were more likely to smoke cigarettes (OR 1.7, 95% CI = 1.0, 2.7]), compared to those in Central Africa. Students from both lower-middle-income (OR 1.9, 95% CI = 1.3, 2.8]) and upper-middle-income countries (OR 3.1, 95% CI [1.9, 5.1]) had higher odds of smoking cigarettes than those in low-income countries. Students from countries that are dominated by Islam and Hinduism were more likely to smoke cigarettes (OR 3.2 [95% CI = 1.6, 6.7]) than students from countries that are dominated by Christianity (Table 3).

Furthermore, students who had parents who used any tobacco products were more likely to smoke cigarettes (OR 2.8, [95% CI = 2.3, 3.4]) than those who did not. Those who had experienced bullying (OR 3.0, [95% CI = 2.5, 3.6]) as well as feelings of sadness and hopelessness [OR 2.4, 95% CI = 2.0, 2.9]) were more likely to smoke cigarettes than those who had not. Those who had missed classes without permission were also more likely to smoke cigarettes (OR 1.7, [95% CI = 1.5, 2.0]) than those who had not (Table 3).

For marijuana use, being a female was associated with lower odds of using marijuana (OR 0.6, [95% CI = 0.5, 0.9]) compared to being a male. Residing in West Africa was associated with higher odds of using marijuana (OR 3.8, [95% CI = 1.7, 8.4]) than in Central Africa. Similarly, adolescents from upper-middle-income countries (OR 5.7, [95% CI = 2.3, 13.9]) had higher odds of using marijuana than those in low-income countries (Table 3).

Having one close friend (OR 0.6, [95% CI = 0.4, 0.9]) or at least two close friends (OR 0.6, [95% CI = 0.4, 1.0]) was associated with higher odds of using marijuana compared to having no close friends. Students who had parents/guardians who used any tobacco products were more likely to use marijuana compared to those who did not have such parents. Those who reported being bullied (OR 2.6, [95% CI = (2.0, 3.4]) and being sad and hopeless (OR 3.1, [95% CI = 2.3, 4.2]) were more likely to be current marijuana users than those who did not report being bullied and being sad and hopeless, respectively. Lastly, those who missed school without permission were more likely to use marijuana (OR 2.4, [95% CI = 1.8, 3.3]) than those who did not (Table 3).

For the dual use of the substances, similar to the single-use, two location variables, residing in West Africa and countries in higher World Bank income groups, were associated with the increased odds of the dual use of all three substances (alcohol and cigarettes, alcohol and marijuana, and cigarettes and marijuana). Similarly, the interpersonal factors: having smoking parents/guardians, being bullied, experiencing sadness and hopelessness, and missing school without permission increased the odds of dual use of any of these substances (see Table 4).

**Table 4.** Logistic regression results of the factors associated with the current dual use of different substance.

| Variables | Dual Alcohol & Cigarettes<br>Odds Ratio (95% CI) | Dual Alcohol & Marijuana<br>Odds Ratio (95% CI) | Dual Cigarettes & Marijuana<br>Odds Ratio (95% CI) |
| --- | --- | --- | --- |
| Age | 1.0 (0.8, 1.2)<br>[0.879] | 1.0 (0.9, 1.2)<br>[0.646] | 1.0 (0.9, 1.2)<br>[0.840] |
| Female (Ref: Male) | 0.8 (0.6, 1.0)<br>[0.088] | 0.9 (0.6, 1.5)<br>[0.795] | 0.7 (0.4, 1.1)<br>[0.137] |
| Education (Ref: Primary) |  |  |  |
| Secondary | 0.9 (0.6, 1.4)<br>[0.570] | 0.5 (0.2, 1.4)<br>[0.189] | 0.4 (0.1, 1.0)<br>[0.056] |
| Africa Region (Ref: East Africa) |  |  |  |
| North Africa | X<br>X | X<br>X | 3.6 (0.3, 39.5)<br>[0.295] |
| Southern Africa | 0.9 (0.6, 1.3)<br>[0.510] | 1.2 (0.3, 4.9)<br>[0.840] | 0.3 (0.0, 3.0)<br>[0.274] |
| West Africa | 2.9 (1.5, 5.9)<br>[0.002] | 4.4 (1.7, 11.7)<br>[0.003] | 6.1 (1.8, 21.2)<br>[0.004] |
| World Bank income group (Ref: Low Income) |  |  |  |
| Lower middle income | 1.9 (1.2, 3.0)<br>[0.010] | 3.8 (0.5, 31.2)<br>[0.216] | 1.3 (0.6, 28.5)<br>[0.858] |
| Upper middle income | 5.8 (3.4, 9.9)<br>[0.000] | 8.8 (2.8, 27.7)<br>[0.000] | 19.3 (4.7, 79.0)<br>[0.000] |
| Religion (Ref: Christian) |  |  |  |
| Muslim & Hindu | 2.4 (0.9, 6.4)<br>[0.076] | 0.6 (0.3, 1.0)<br>[0.061] | 0.8 (0.4, 1.3)<br>[0.319] |
| Having close friends (Ref: None) |  |  |  |
| One | 1.0 (0.6, 1.8) | 0.7 (0.3, 1.7) | 1.0 (0.5, 2.1) |
|  | [0.741] | [0.430] | [0.953] |
| At least two | 1.3 (0.8, 2.3) | 1.4 (0.6, 3.2) | 1.3 (0.7, 2.7) |
|  | [0.285] | [0.387] | [0.407] |
| Parental smoking (Ref: No) | 4.1 (2.8, 6.1) | 6.5 (4.0, 10.6) | 5.2 (3.2, 8.5) |
|  | [0.000] | [0.000] | [0.000] |
| Bullying (Ref: No) | 3.6 (2.6, 5.1) | 3.7 (2.3, 5.9) | 4.3 (2.6, 7.1) |
|  | [0.000] | [0.000] | [0.000] |
| Sad & hopeless (Ref: No) | 2.4 (1.8, 3.3) | 2.6 (1.7, 4.2) | 3.9 (2.4, 6.3) |
|  | [0.000] | [0.000] | [0.000] |
| Missed school (Ref: No) | 2.0 (1.5, 2.5) | 2.3 (1.3, 3.9) | 2.0 (1.2, 3.1) |
|  | [0.000] | [0.003] | [0.004] |
| Observations | 22,775 | 12,233 | 18,214 |

*Notes:* The p-values are reported in square brackets. The regression results were weighted, and the standard errors have been adjusted for clustering. The survey year fixed effects have been included in all three regressions. X indicates that the data was unavailable. Urbanization was dropped due to multicollinearity.

## DISCUSSION

We found that the overall prevalence of alcohol use (10.6%) was the highest, followed by cigarette smoking (6.9%) and then marijuana use (3.8%). The co-consumption of alcohol and cigarettes was more prevalent than that of alcohol and marijuana and cigarettes and marijuana. In terms of factors associated, some factors had a significant association with particular substances, but not all. Irrespective of substance type, residing in West Africa and upper-middle-income countries, having parents that use tobacco products, experiencing bullying, feeling hopeless and sad, and missing school without permission were risk factors of the current use of alcohol, cigarettes, and marijuana. Being a girl was a protective factor against the use of all substances. Predominant country-level religion and education level were significant only for cigarettes while having friends was positively associated with the use of alcohol but negatively associated with marijuana.

The significantly high prevalence of alcohol drinking compared to cigarettes and marijuana may be explained by the fact that alcohol use is generally more accepted than tobacco or marijuana use in many cultures.^2^ In many countries, alcohol is also more available, accessible, and cheaper than other products.^3^ Marijuana has the lowest prevalence among the three substances potentially because marijuana has long been considered illegal in all African countries and, therefore, might be the least socially acceptable substance. Some African countries, such as Lesotho and Zimbabwe, have legalized medical marijuana use (in 2017 and 2018, respectively), but recreational use remains illegal.^16^ Our finding that alcohol use is more common than cigarette smoking or marijuana use, is in line with other studies in Africa.^7^ Our alcohol prevalence estimates align with previous studies that have utilized the GSHS.^2 9^

Despite using a different number of African countries, our cigarette smoking prevalence estimates are not significantly different from the estimates in the study of 22 African countries based on the Global Youth Tobacco Survey (GYTS) data.^17^ For marijuana prevalence, our estimates are in line with the study of 8 sub-Saharan countries that found the overall prevalence was between 4% and 5%,^5^ with Seychelles having the highest marijuana use prevalence.^5 8^ Our estimates for alcohol and marijuana highlight the lack of data on monitoring marijuana and alcohol use among adolescents in African countries. Currently, the GSHS is the available primary data source for providing information on alcohol and marijuana use among young people in the WHO Member states. As shown in Supplementary Table 1, out of 54 African countries, only 13 and 15 countries, respectively, have publicly available datasets on alcohol and marijuana use.

Our prevalence estimates of the co-consumption of substances suggest that adolescents who smoke and drink have higher chances of developing addiction to these products, have lower chances of quitting, and are likely to continue use in the long term.^18^ Those who co-consume cigarettes and alcohol also have an increased risk of having difficulties at school, delinquency,^19^ and emotional development for those in low socioeconomic status,^20^ than those who do not co-consume these products. Co-consumption of alcohol and marijuana substantially increases the likelihood of comorbid substance use and experiencing mental health disorders.^21^

In terms of the factors, experiencing bullying, sadness, and hopelessness as risk factors of substance use were also evident in 8 other sub-Saharan African countries.^8^ Nonetheless, unlike our study, these factors were only significant for alcohol use, not for cigarettes or marijuana. Unlike evidence from 73 LMICs that a country’s wealth matters only for marijuana smoking, ours suggests that country-level income matters significantly for the prevalence of use of all substances in Africa. Furthermore, consistent with other studies,^5 8^ being a female was a protective factor of being a cigarette or marijuana smoker. The predominant religion (i.e., Islam) at the country level was a risk factor for cigarette smoking only. This is in contrast with evidence from 73 low- and -middle-income countries,^2^ which suggested that the Islamic religion was a protective factor against alcohol use.

Given that young people are mainly sensitive to increases in the prices (hence taxes) of cigarettes,^22^ and alcohol,^23^ countries need to increase their excise taxes to minimize the prevalence rates. This could be complemented by strengthening restrictions on the availability of the products, especially alcohol, which is very prevalent among African school-going adolescents. As recommended by WHO,^3^ comprehensive restrictions on alcohol advertising, promotion, and sponsorship should be enforced.

Although this study appears to be the first with a larger sample size using the GSHS data and broader coverage of African countries, it has limitations. First, the fact that the available GSHS datasets are outdated means that they explain the situation as it was in those survey years, not currently. Nonetheless, given the lack of data, this provides us with insights about substance use in Africa. Similarly, given the institutions, such as cultural norms, are often persistent and do not change quickly, these datasets still shed light on the current situation. Second, the results cannot be generalized to all adolescents, especially those not in school, since GSHS does not include those who do not attend school or were absent during the survey. Third, our logit regression models do not include all factors that theoretically and empirically are associated with these substances. For instance, price, pocket money, and media-related factors have not been included. The prices play a significant role in determining the use of alcohol,^23^ cigarettes,^23^ or marijuana.^24^ Thus, for future research, all these limitations could be addressed to improve the robustness of the results.

In conclusion, our results on the prevalence and determinants each shed light on policy directions. First, comprehensive data on substance use among adolescents in and out of school in Africa is needed for effective monitoring and policy-making. Second, to minimize substance use, primary and secondary schools should develop mentorship policies/programs that reduce the rates of school absenteeism, positively impact students’ behaviour, and reduce bullying among them.

## Data Availability

All the datasets are publicly available

https://www.who.int/teams/noncommunicable-diseases/surveillance/systems-tools/global-school-based-student-health-survey

## ACKNOWLEDGMENTS

This study was conducted as part of the Data on Youth and Tobacco in Africa (DaYTA) program. This program seeks to empower stakeholders to make timely, data-driven decisions by using evidence to inform policy and advance tobacco control efforts in sub-Saharan Africa. The program achieves this by addressing data gaps related to tobacco use among 10-17-year-olds in Kenya, Nigeria, and the Democratic Republic of Congo. The DaYTA program is led by Development Gateway: an IREX Venture (DG), a global non-profit organization specializing in data for development.

## CONTRIBUTORS

RP, TGA, and NDM conceptualized the study. RP conducted the data analysis and wrote the first draft of the manuscript. TGA, JDT and NDM contributed to the drafting and revision of the manuscript.

## PATIENT CONSENT FOR PUBLICATION

Not required.

## DATA SHARING STATEMENT

All datasets are publicly available.

