## Supplementary Information for "The prevalence and factors associated with alcohol, cigarette, and marijuana use among adolescents in 25 African countries: evidence from Global School-Based Health Surveys"

**Supplementary Table 1: Latest data availability on the use of cigarettes, alcohol, and marijuana in African GSHS among adolescents, 2003 - 2017**

| **Location** | **Cigarettes** | **Alcohol** | **Marijuana** |
| --- | --- | --- | --- |
| **East Africa** |  |  |  |
| Djibouti (2007) | ✔ | X | X |
| Kenya (2003) | ✔ | ✔ | X |
| Mauritius (2017) | ✔ | X | ✔ |
| Seychelles (2015) | ✔ | ✔ | ✔ |
| Sudan (2012) | ✔ | X | X |
| Tanzania (2014) | ✔ | ✔ | ✔ |
| Uganda (2003) | ✔ | ✔ | X |
| **Total available datasets** | **7** | **4** | **3** |
| **North Africa** |  |  |  |
| Algeria (2011) | ✔ | X | ✔ |
| Egypt (2011) | ✔ | X | X |
| Libya (2007) | ✔ | X | X |
| Mauritania (2010) | ✔ | X | ✔ |
| Morocco (2016) | ✔ | X | ✔ |
| Tunisia (2008) | ✔ | X | X |
| **Total available datasets** | **6** | **0** | **3** |
| **Southern Africa** |  |  |  |
| Botswana (2005) | ✔ | ✔ | X |
| Eswatini (2013) | X | X | ✔ |
| Malawi (2009) | ✔ | ✔ | X |
| Mozambique (2015) | ✔ | ✔ | ✔ |
| Namibia (2013) | ✔ | ✔ | ✔ |
| Zambia (2004) | X | ✔ | X |
| Zimbabwe (2003) | ✔ | ✔ | X |
| **Total available datasets** | **5** | **6** | **3** |
| **West Africa** |  |  |  |
| Benin (2016) | ✔ | ✔ | ✔ |
| Ghana (2017) | ✔ | ✔ | ✔ |
| Liberia (2008) | ✔ | ✔ | ✔ |
| Senegal (2005) | ✔ | ✔ | X |
| Sierra Leone (2017) | X | ✔ | ✔ |
| **Regional Total** | **4** | **5** | **4** |
| **Total available GSHS in Africa** | **22** | **15** | **13** |

*Notes:* ✔ indicates data availability, and X indicates data unavailability.

**Supplementary Table 2. Prevalence of alcohol, cigarettes, and marijuana use: by region, income group, and country**

|  | **Alcohol use** | | **Cigarette smoking** | | | **Marijuana** | | |
| --- | --- | --- | --- | --- | --- | --- | --- | --- |
|  | **Boys (95% CI)** | **Girls (95% CI)** | **Boys (95% CI)** | | **Girls (95% CI)** | **Boys (95% CI)** |  | **Girls (95% CI)** |
| **East Africa** |  | | | | | | |  |
| Djibouti (2007) | X | X | 8.6% (6.3, 11.5) | 2.6% (1.5, 4.6) | | X |  | X |
| Kenya (2003) | 14.7% (10.2, 20.8) | 11.1% (7.1, 16.9) | 19.3% (14.7, 24.9) | 13.1% (9.0, 18.5) | | X |  | X |
| Mauritius (2017) | 24.6% (20.7, 29.0) | 22.0% (17.1, 28.0) | 22.4% (18.2, 27.2) | 11.1% (7.7, 15.7) | | 9.6% 7.8, 11.9) |  | 1.4% (0.8, 2.3) |
| Seychelles (2015) | 47.1% (43.0, 51.2) | 48.1% (44.6, 51.7) | 24.5% (21.3, 27.9) | 14.8% (12.4, 17.5) | | 13.3% (10.6, 16.7) |  | 5.4% (4.0, 7.2) |
| Sudan (2012) | X | X | 8.8% (6.9, 11.2) | 4.5% (2.7, 7.5) | | X |  | X |
| Tanzania (2014) | 4.6% (3.4, 6.2) | 4.1% (2.8, 5.9) | 5.4% (3.9, 7.5) | 3.6% (2.5, 5.2) | | 2.3% (1.4, .9) |  | 2.6% (1.9, 3.6) |
| Uganda (2003) | 9.2% (7.5, 11.1) | 9.2% (7.5, 11.1) | 6.8% (5.2, 8.8) | 2.9% (1.9, 4.5) | | X |  | X |
| **Regional prevalence** | **8.3% (6.7, 10.2)** | **6.7% (5.1, 8.6)** | **9.7% (8.1, 11.5)** | **6.1% (4.8, 7.6)** | | **2.5% (1.6, 3.9)** |  | **2.6% (1.9, 3.6)** |
| **North Africa** |  |  |  |  | |  |  |  |
| Algeria (2011) | X | X | 18.5% (15.1, 22.6) | 1.3% (0.9, 2.1) | | 4.0% (2.6, 5.9) |  | 0.4% (0.2, 1.0) |
| Egypt (2011) | X | X | 8.4% (4.8, 14.4) | 1.7% (0.8, 3.2) | | X |  | X |
| Libya (2007) | X | X | 6.7% (5.4, 8.3) | 1.5% (0.8, 2.6) | | X |  | X |
| Mauritania (2010) | X | X | 17.9% (14.3, 22.1) | 16.4% (11.9, 22.2) | | 7.0% (3.8, 12.5) |  | 6.3% (3.2, 12.2) |
| Morocco (2016) | X | X | 8.4% (7.1, 9.8) | 2.8% (2.1, 3.7) | | 8.0% (6.2, 10.4) |  | 2.2% (1.4, 3.4) |
| Tunisia (2008) | X | X | 13.9% (10.7, 17.9) | 3.0% (1.9, 4.8) | | X |  | X |
| **Regional prevalence** | X | X | **10.7% (8.3, 13.6)** | **2.0% (1.5, 2.6)** | | **6.1% (4.9, 7.6)** |  | **1.3% (0.9, 2.0)** |
| **Southern Africa** |  |  |  |  | |  |  |  |
| Botswana (2005) | 24.1% (21.2, 27.2) | 21.1% (18.0, 24.5) | 10.9% (9.6, 12.4) | 4.8% (3.6, 6.5) | | X |  | X |
| Eswatini (2013) | X | X | X | X | | 5.2% (3.8, 7.1) |  | 1.4% (0.8, 3.6) |
| Malawi (2009) | 5.3% (2.9, 9.4) | 2.4% (0.8, 6.4) | 5.9% (4.0, 8.5) | 3.5% (2.0, 6.2) | | X |  | X |
| Mozambique (2015) | 10.9% (7.4, 15.7) | 10.6% (6.7, 16.4) | 1.2% (0.5, 3.0) | 2.9% (1.1, 7.4) | | 2.3% (1.0, 5.1) |  | 0.5% (0.1, 2.4) |
| Namibia (2013) | 29.7% (25.7, 34.1) | 24.7% (22.8, 26.7) | 12.0% (9.8, 14.7) | 6.0% (4.2, 8.5) | | 6.8% (5.1, 9.0) |  | 3.7% (2.3, 5.8) |
| Zambia (2004) | 24.5% (20.8, 28.6) | 28.9% (26.1, 31.8) | X | X | | X |  | X |
| Zimbabwe (2003) | 17.6% (14.6, 21.0) | 10.9% (8.2, 14.4) | 12.2% (10.3, 14.4) | 7.4% (5.7, 9.5) | | X |  | X |
| **Regional Prevalence** | **13.3% (11.4, 15.4)** | **12.0% (10.1, 14.1)** | **5.8% (4.6, 7.2)** | **4.1% (3.0, 5.6)** | | **3.2% (0.3, 0.9)** |  | **1.3% (0.7, 2.2)** |
| **West Africa** |  |  |  |  | |  |  |  |
| Benin (2016) | 40.9% (35.3, 46.8) | 38.2% (32.9, 43.9) | 5.6% (3.5, 8.9) | 1.3% (0.6, 2.8) | | 1.3% (0.5, 3.3) |  | 0.2% (0.0, 1.2) |
| Ghana (2017) | 16.1% (13.8, 18.6) | 13.1% (9.9, 17.2) | 8.9% (6.4, 12.2) | 7.8% (4.3, 13.6) | | 6.0% (4.5, 8.0) |  | 8.4% (5.2, 13.2) |
| Liberia (2008) | 21.6% (16.9, 27.1) | 13.7% (10.3, 18.0) | 7.1% (4.8, 10.4) | 5.9% (4.2, 8.2) | | 7.2% (4.6, 11.2) |  | 4.3% (2.3, 7.9) |
| Senegal (2005) | 7.0% (3.8, 12.6) | 4.0% (2.3, 6.8) | 12.2% (6.5, 21.6) | 3.2% (1.1, 9.3) | | X |  | X |
| Sierra Leone (2017) | 14.8% (10.7, 20.1) | 8.6% (6.2, 11.8) | X | X | | 5.6% (3.5, 8.6) |  | 2.8% (1.3, 5.8) |
| **Regional Prevalence** | **17.4% (15.6, 19.5)** | **12.9% (10.7, 15.5)** | **9.2% (6.8, 12.3)** | **6.1% (3.7, 9.8)** | | **5.0% (4.0, 6.4)** |  | **6.7% (4.3, 10.1)** |
| **World Bank income group** |  |  |  |  | |  |  |  |
| Upper middle-income | 27.1% (24.9, 29.4) | 23.4% (21.7, 25.2) | 9.0% (7.8, 10.2) | 3.7% (2.4, 5.3) | | 7.9% (6.6, 9.3) |  | 3.1% (2.1, 4.5) |
| Lower middle income | 11.8% (10.3, 13.4) | 9.0% (7.6, 10.7) | 10.4% (8.8, 12.3) | 4.0% (3.3, 4.8) | | 4.6% (3.8, 5.5) |  | 2.8% (2.2, 1.1) |
| Low income | 10.1% (8.4, 12.0) | 7.4% (5.9, 9.3) | 6.0% (4.9, 7.3) | 3.7% (2.7, 4.9) | | 3.6% (2.4, 5.3) |  | 1.5% (0.9, 2.5) |
| **Overall prevalence** | **11.9% (10.7, 13.2)** | **9.2% (8.1, 10.5)** | **9.8% (8.4, 11.2)** | **3.9% (3.3, 4.6)** | | **4.6% (3.9, 5.3)** |  | **2.7% (2.2, 3.4)** |

*Notes:* X indicates data unavailability

**Supplementary Table 3. Prevalence of dual use of the substances by region, income group, and country**

|  | **Dual alcohol & cigarette use** | | **Dual alcohol & marijuana smoking** | | | **Dual cigarette & marijuana smoking** | | |
| --- | --- | --- | --- | --- | --- | --- | --- | --- |
|  | **Boys (95% CI)** | **Girls (95% CI)** | **Boys (95% CI)** | | **Girls (95% CI)** | **Boys (95% CI)** |  | **Girls (95% CI)** |
| **East Africa** |  | | | | | | |  |
| Djibouti (2007) | X | X | X | X | | X |  | X |
| Kenya (2003) | 7.8% (4.5, 13.3) | 7.8% (4.5, 13.3) | X | X | | X |  | X |
| Mauritius (2017) | 12.6% (10.0, 15.9) | 5.8% (3.0, 11.0) | 6.4% (5.2, 7.7) | 1.1% (0.6, 2.0) | | 7.0% (5.5, 8.8) |  | 1.0% (0.6, 1.8) |
| Seychelles (2015) | 17.5% (14.7, 20.8) | 13.2% (10.9, 15.8) | 9.2% (7.2, 11.8) | 4.1% (3.0, 5.7) | | 9.0% (7.1, 11.4) |  | 3.5% (2.5, 4.9) |
| Sudan (2012) | X | X | X | X | | X |  | X |
| Tanzania (2014) | 1.4% (0.70, 2.6) | 1.2% (0.7, 2.3) | 1.1% (0.6, 1.9) | 1.3% (0.8, 2.1) | | 0.5% (0.3, 1.4) |  | 1.0% (0.6, 1.7) |
| Uganda (2003) | 2.9% (2.1, 4.0) | 1.2% (0.8, 1.9) | X | X | | X |  | X |
| **Regional prevalence** | **8.3% (6.7, 10.2)** | **2.3% (1.4, 3.6)** | **0.8% (0.5, 1.4)** | **0.9% (0.5, 1.4)** | | **0.4% (0.2, 0.7)** |  | **0.6% (0.4, 1.0)** |
| **North Africa** |  |  |  |  | |  |  |  |
| Algeria (2011) | X | X | X | X | | 2.4% (1.4, 4.3) |  | 0.1% (0.0, 0.3) |
| Egypt (2011) | X | X | X | X | | X |  | X |
| Libya (2007) | X | X | X | X | | X |  | X |
| Mauritania (2010) | X | X | X | X | | 4.3% (2.3, 7.8) |  | 4.3% (1.9, 9.1) |
| Morocco (2016) | X | X | X | X | | 3.1% (2.4, 4.0) |  | 0.6% (0.3, 1.1) |
| Tunisia (2008) | X | X | X | X | | X |  | X |
| **Regional prevalence** | X | X | X | X | | **1.3% (0.9, 1.7)** |  | **0.2% (0.1, 0.3)** |
| **Southern Africa** |  |  |  |  | |  |  |  |
| Botswana (2005) | 6.8% (5.7, 8.1) | 3.1% (2.4, 4.0) | X | X | | X |  | X |
| Eswatini (2013) | X | X | X | X | | X |  | X |
| Malawi (2009) | 5.3% (2.9, 9.4) | 0.8% (0.2, 2.4) | X | X | | X |  | X |
| Mozambique (2015) | 0.6% (0.2, 1.9) | 1.2% (0.5, 3.1) | 1.8% (0.7, 4.7) | 0.1% (0.0, 0.7) | | 0.6% (0.2, 2.2) |  | X |
| Namibia (2013) | 8.3% (6.6, 10.3) | 3.9% (2.8, 5.6) | 4.2% (3.1, 5.7) | 2.1% (1.3, 3.3) | | 3.3% (2.4, 4.6) |  | 2.0% (1.1, 3.5) |
| Zambia (2004) | X | X | X | X | | X |  | X |
| Zimbabwe (2003) | 5.9% (4.4, 7.8) | 3.0% (2.1, 4.2) | X | X | | X |  | X |
| **Regional Prevalence** | **2.5% (1.9, 3.2)** | **1.5% (1.0, 2.1)** | **0.9% (0.4, 1.7)** | **0.2% (0.1, 0.4)** | | **0.5% (0.3, 0.9)** |  | **0.2% (0.1, 0.4)** |
| **West Africa** |  |  |  |  | |  |  |  |
| Benin (2016) | 4.9% (3.1, 7.7) | 1.3% (0.6, 2.7) | 1.0% (0.3, 3.2) | 0.2% (0.0, 1.2) | | 0.5% (0.2, 1.7) |  | 0.2% (0.0, 1.2) |
| Ghana (2017) | 4.8% (3.4, 6.6) | 4.7% (2.6, 8.5) | 4.2% (3.1, 5.7) | 4.0% (2.6, 6.1) | | 2.8% (1.6, 5.0) |  | 3.5% (1.9, 6.3) |
| Liberia (2008) | 3.1% (1.4, 6.7) | 2.2% (1.2, 4.2) | 3.2% (1.9, 5.4) | 1.6% (0.7, 3.5) | | 2.3% (1.1, 4.7) |  | 2.0% (1.0, 3.9) |
| Senegal (2005) | 4.2% (2.0, 8.6) | 0.4% (0.2, 1.1) | X | X | | X |  | X |
| Sierra Leone (2017) | X | X | X | X | | X |  | X |
| **Regional Prevalence** | **4.3% (3.3, 5.6)** | **3.1% (1.8, 5.3)** | **2.7% (2.0, 3.5)** | **2.6% (1.8, 3.9)** | | **1.6% (0.9, 2.7)** |  | **2.2% (1.2, 3.8)** |
| **World Bank income group** |  |  |  |  | |  |  |  |
| Upper middle-income | 2.8% (2.3, 3.5) | 1.5% (1.1, 1.9) | 3.6% (3.0, 4.4) | 1.3% (0.9, 2.0) | | 1.0% (0.7, 1.3) |  | 0.4% (2.1, 4.5) |
| Lower middle income | 1.8% (1.4, 2.4) | 1.3% (0.9, 1.8) | 0.9% (0.7, 1.2) | 0.9% (0.6, 1.2) | | 1.1% (0.9, 1.5) |  | 0.6% (0.4, 0.9) |
| Low income | 1.2% (0.9, 1.7) | 0.7% (0.4, 1.2) | 0.3% (0.2, 0.5) | 3.7% (2.7, 4.9) | | 0.2% (0.1, 0.5) |  | 0.1% (0.0, 0.2) |

*Notes:* X indicates data unavailability.

**Supplementary Table 4: Sensitivity analysis (robustness checks) of the prevalence of different substances by gender, African region, and World Bank income group (2010 – 2017)**

|  | **Alcohol (95% CI)** | **Cigarettes (95% CI)** | **Marijuana (95% CI)** | **Dual alcohol & cigarettes (95% CI)** | **Dual alcohol & marijuana (95% CI)** | **Dual cigarettes & marijuana (95% CI)** |
| --- | --- | --- | --- | --- | --- | --- |
| **African region** |  |  |  |  |  |  |
| East Africa | 5.0% (3.9, 6.5) | 5.4% (4.4, 6.5) | 2.7% (2.0, 3.7) | 1.3% (0.9, 1.9) | 1.4% (1.0, 1.9) | 0.8% (0.6, 1.1) |
| North Africa | 🞩 | 6.5% (5.0, 8.3) | 3.9% (3.1, 4.8) | 🞩 | 🞩 | 0.8% (0.6, 1.1) |
| Southern Africa | 13.5% (10.5, 17.3) | 3.5% (2.2, 5.4) | 2.2% (1.5, 3.1) | 1.8% (1.2, 2.6) | 1.3% (0.7, 2.2) | 0.7% (0.4, 1.2) |
| West Africa | 18.1% (16.2, 20.3) | 7.7% (5.5, 10.7) | 6.0% (4.4, 8.1) | 4.7% (3.6, 6.1) | 3.4% (2.5, 4.5) | 2.4% (1.5, 3.8) |
| **World Bank Income Group** |  |  |  |  |  |  |
| Upper middle income | 26.5% (24.8, 28.2) | 11.5% (10.0, 13.1) | 5.2% (4.3, 6.4) | 7.3% (6.4, 8.3) | 3.3% (2.7, 4.1) | 3.1% (2.4, 3.9) |
| Lower middle income | 9.3% (8.0, 10.9) | 6.2% (5.2, 7.5) | 3.8% (3.2, 4.5) | 0.9% (0.7, 1.2) | 1.1% (0.9, 1.4) | 1.1% (0.9, 1.3) |
| Low income | 11.5% (9.1, 14.5) | 5.0% (3.8, 6.6) | 2.6% (1.8, 3.6) | 0.5% (0.3, 0.8) | 1.5% (0.9, 2.3) | 0.3% (0.1, 0.5) |
| **Gender** |  |  |  |  |  |  |
| Boys | 11.6% (10.2, 13.3) | 9.0% (7.3, 10.9) | 4.6% (3.9, 5.3) | 1.1% (0.8, 1.4) | 1.3% (1.0, 1.6) | 1.3% (1.0, 1.6) |
| Girls | 8.9% (7.4, 10.5) | 3.1% (2.5, 3.9) | 2.7% (2.2, 3.4) | 0.9% (0.6, 1.2) | 1.1% (0.8, 1.4) | 0.7% (0.5, 1.0) |
| **Overall prevalence** | **10.3% (9.0, 11.7)** | **6.2% (5.2, 7.3)** | **3.8% (3.2, 4.4)** | **1.0%(0.8, 1.2)** | **1.2% (1.0, 1.5)** | **1.0%(0.8, 1.2)** |

*Notes:* 🞩 indicates data unavailability.
